# A protocol to assess antipsychotics prescribing and physical health monitoring in children and young people: a cohort study using primary care data from QResearch

**DOI:** 10.1101/2024.07.18.24310326

**Authors:** Yana Vinogradova, Ruth H Jack, Vibhore Prasad, Carol Coupland, Richard Morriss, Chris Hollis

## Abstract

Antipsychotic medicines are prescribed to children and young people (CYP) with mental health conditions such as schizophrenia, bipolar disorder, behavioural disorders, autism spectrum disorder and tics. The physical side effects of these medicines require monitoring, and this responsibility can be transferred to primary care. This study will describe the trends in antipsychotic prescribing in CYP, identify the mental health conditions, symptoms and comorbidities associated with the prescriptions, and determine the extent of physical health monitoring in primary care both for CYP prescribed antipsychotics and those with mental health conditions without antipsychotic prescriptions. The study will use data from QResearch, a large, anonymised, UK primary care electronic health record database. Records will be linked to secondary care data from hospital episode statistics and socioeconomic deprivation information. People aged 5 to 17 years, who are registered with GPs in England contributing to QResearch between 1 January 2006 and 31 July 2021 will be included. Incidence and prevalence rates will be calculated over the study period, and Poisson regression will be used to estimate incidence rate ratios. Results from this study will identify where improvements to clinical practice or to treatment guidelines may be needed.

## Introduction

Antipsychotic medicines can be prescribed to children and adolescents with particular mental health issues. These include severe conditions, such as schizophrenia,^1^ as well a range of behavioural disorders, learning difficulties, autism spectrum disorder and tics.^2^ Patients often require long-term treatment, so the drugs may be prescribed for months or years. It is known that antipsychotics have side effects, which include substantial weight gain and increased risk of developing type 2 diabetes.^3, 4^ It is also known that young people are at greater risk of experiencing these side effects because their metabolisms may require higher dosages.^4^ There is, however, limited evidence about the health monitoring of people prescribed antipsychotics, and most studies to date have been short-term and have focused on adult patients.^5 6^

Because of the greater risks of side effects from antipsychotics usage in the younger population, the National Institute of Health and Clinical Excellence (NICE) has recommended regular physical health monitoring for some younger antipsychotic users.^1, 7^ However, it is currently not known how widely or closely these recommendations are followed. We shall therefore investigate the recorded physical health monitoring for CYP with diagnoses of mental health conditions which have led to, or could lead to, the prescribing of an antipsychotic.

Little is known about the extent of or reasons for prescribing of antipsychotics to younger people, or about the procedures followed when this is done. This study will investigate antipsychotic prescribing to children and adolescents in real primary care settings in the UK. We will include recorded reasons for prescribing, and how long patients were prescribed drugs for. The data used will come from a national database of GP records, which has been shown to be representative of the general population,^8^ so all types of antipsychotics used in the UK – and all population groups and patient characteristics – will be included.

This study will clarify levels of prescribing and use of antipsychotics for the treatment of mental health issues in children and adolescents, the reasons for prescribing and procedures followed, the background levels of health monitoring of younger people with mental health issues and whether these levels change when patients have been prescribed these drugs.

## Methods

### Patient or user group involvement

The research question was initially presented to the National Institute for Health Research MindTech HealthTech Research Centre Involvement Team, a group of individuals with lived experience of mental health conditions. Members of this group will attend project meetings and be involved in discussions throughout the study. They will also provide advice on dissemination activities.

### Study design and data sources

We will examine incidence and prevalence of antidepressant prescribing in a cohort of CYP in England using a large primary care database (QResearch) linked to HES admitted patient care and outpatient data. The QResearch database includes health records of over 35□million people from general practices across the UK which record data using the Egton Medical Information Systems (EMIS) medical records computer system. The information recorded includes personal characteristics, clinical diagnoses, symptoms and prescribed medicines.

### Study population

We will identify an open cohort of patients aged 5 to 17 years, who are registered with GPs contributing to QResearch between 1 January 2006 and 31 July 2021. Patients’ entry will be the latest date of: installation of the GP’s EMIS computer system plus 1 year; registration with the practice plus 3 years; start of their 5^th^ birthday year; start of study period (1 January 2006). Patients will leave the study population at the earliest of: date of practice leaving QResearch; deregistration from the practice; start of their 18^th^ birthday year; their death; end of study period (31 July 2021).

### Outcomes

We will examine the incidence and prevalence of antipsychotics prescribed overall and the most commonly prescribed individual medicines (aripiprazole, olanzapine, prochlorperazine, quetiapine and risperidone). Having at least one antipsychotic prescription within a year will be counted as an outcome for the time trend analysis.

Indications for those with an incident antipsychotic prescription will be identified using Read and ICD-10 code lists recorded before the prescription date in GP records, HES admission and outpatient data. These are: mental health conditions (psychosis/schizophrenia, bipolar disorder, ASD and Tourette syndrome), mental health symptoms (aggression/disruptive behaviour and self-harm) and mental health comorbidities (anxiety, attention-deficit/hyperactivity disorder (ADHD), depression, eating disorders and learning difficulties). Code lists will come from the QResearch library or developed by the study team. A single mental health condition will be recorded for each person. If multiple mental health conditions are present, these will be prioritised in the following order: psychosis/schizophrenia, bipolar disorder, ASD, Tourette syndrome. Duration of antipsychotics will be determined using prescription information, with gaps of less than 90 days considered a continuous prescription.

Physical health monitoring will be identified using records measuring the heart (systolic/diastolic blood pressure and pulse), anthropometrics (height, weight, body-mass index), lipids (total, high and low density cholesterols), blood glucose (fasting glucose and HbA1c), prolactin, diagnosis of diabetes.

### Covariates

We will examine prevalence and incidence rates of antipsychotics prescribing separately in males and females in two age groups: a younger group aged 5 to 11 years old and an older group aged 12 to 17 years. We will also examine incidence rates and rate ratios by region, Townsend deprivation score quintile^17^ and ethnicity (White, Black, Asian, Other [including Mixed] and Unknown) based on the England and Wales 2001 Census groups.

### Data/statistical analysis

Characteristics of the group prescribed antipsychotics will be described. Incidence and prevalence prescribing rates per 10,000 person-years will be calculated in each year for the four age-sex groups described in the covariates section. For each of the covariate groups, we will calculate incidence rate ratios using Poisson regression, and the median and interquartile range of treatment in months. The median and interquartile range of the length of treatment will also be calculated for the different mental health conditions, symptoms and comorbidities, overall and in the two age groups.

We will calculate the number and proportion of CYP with different physical health monitoring measurements for both those prescribed antipsychotics within a 2-year follow-up period, and those with mental health conditions and no antipsychotic prescriptions. Of those prescribed antipsychotics, the median and interquartile range will be reported for the number of days during follow-up there was an active prescription.

A sensitivity analysis on physical health monitoring, restricting the study period to 2013 to 2021 will be performed to assess the period after the 2013 NICE guidelines on psychosis and schizophrenia in CYP were published.

### Limitations

There are limitations of our study. We will only be able to identify prescribing and health monitoring within primary care records and cannot determine whether secondary or community care providers were providing these services. Only coded records, rather than free text, will be available and so we may miss some instances of physical health monitoring that took place within primary care.

## Conclusion

This study will give a picture of antipsychotic prescribing in CYP in the UK and whether this varies by demographic characteristics. We will also determine which mental health conditions, symptoms and comorbidities are recorded with initial antipsychotic prescriptions, and what physical health monitoring took place within primary care. The findings will identify any required improvements to clinical practice or to treatment guidelines and areas where more detailed studies may be feasible. It will also highlight possible risks for younger antipsychotic users and stimulate further investigation into the scale of these risks.

## Data Availability

To guarantee the confidentiality of patient information, only the authors will have access to the data during the study in accordance with the relevant licence agreements. Access to the QResearch data is according to the information on the QResearch website (www.qresearch.org).

## Ethics and data sharing

The project has been reviewed in accordance with the QResearch agreement with East Midlands—Derby Research Ethics Committee [reference 18/EM/0400] and approved by the QResearch Scientific Committee on 23 September 2020 (N 4154 / 4129). To guarantee the confidentiality of patient information, only the authors will have access to the data during the study in accordance with the relevant licence agreements. Access to the QResearch data is according to the information on the QResearch website (www.qresearch.org).

## Acknowledgments

The study was funded by NIHR Nottingham Biomedical Research Centre (RC48LF). This work will use data provided by patients and collected by the NHS as part of their care and support. We acknowledge the contribution of EMIS practices who contribute to the QResearch® database and EMIS Health and the Universities of Nottingham and Oxford for expertise in establishing, developing and supporting the QResearch database. The HES data that will be used in this analysis are re-used by permission from the NHS England who retain the copyright. We thank the Office for National Statistics (ONS) for providing mortality data. We would like to acknowledge Julia Hippisley-Cox’s contribution to the development of the protocol, funding application and data specification.

CH and RM are supported by the National Institute for Health Research (NIHR) Nottingham Biomedical Research Centre and NIHR MindTech HealthTech Research Centre. RM is additionally supported by the NIHR Applied Research Collaboration East Midlands (ARC EM). VP received salary funding via King’s College London from the NIHR academic clinical lecturer scheme, University of Nottingham, the NIHR East Midlands Scholarship scheme (hosted by NHS Nottingham and Nottinghamshire and the University of Nottingham) and NIHR Senior Clinical and Practitioner Research Award.

